# Genetic predictors of clonal contraction and hematologic response in *IDH* mutant myeloid malignancies

**DOI:** 10.64898/2026.09.03.26361825

**Authors:** Amy Song, Yi Chen, Sunil Iyer, Sebastian Fernando, Varun Sudunagunta, Edna M. Stewart, Rong Deng, Viviana Scoca, Jane J. Xu, Benjamin L. May, Alina Varabyeva, Susan J Hsiao, Christopher Freeman, Mahesh Mansukhani, Andrew Lipsky, Nicole Lamanna, Todd Rosenblat, Monica Kasbekar, Joseph Jurcic, Brian Chernak, Aaron D. Viny

## Abstract

IDH inhibitors promote differentiation and hematological improvement in myeloid malignancies by reversing epigenetic dysregulation. However, molecular predictors of response remain poorly defined. We retrospectively analyzed 79 patients with *IDH*-mutated myeloid malignancies treated with either ivosidenib (IDH1; n=36) or enasidenib (IDH2; n=43). Hematologic benefit was assessed using a composite complete hematologic response (CHR), defined by neutrophil and platelet recovery, and transfusion independence. Targeted sequencing was performed at baseline and at best hematologic response. Associations between co-mutations, hematologic response, and treatment duration were evaluated using multivariable models. Overall, 43% of patients achieved CHR, with similar rates between inhibitors. Ivosidenib-treated patients achieving CHR demonstrated a significant reduction in *IDH1* VAF (*p*=0.010), which was not observed in enasidenib-treated CHR patients. Within the CHR cohort, ivosidenib patients had a greater reduction in *IDH* VAF relative to enasidenib patients (mean VAF change -16.74% vs - 0.05%, *p=*0.034). Ivosidenib responders demonstrated a trend toward more durable treatment duration compared with enasidenib responders (p = 0.0908). *RUNX1* and *BCOR* mutations were significantly enriched among patients with incomplete hematologic response and were independently associated with shorter treatment duration after adjustment for clinical factors (*RUNX1*: HR 2.53, *p*=0.0024; *BCOR*: HR 2.31, *p*=0.0058). IDH1 inhibition was associated with clonal contraction and durable hematologic benefit, a pattern not observed with IDH2 inhibition. Co-mutations in *RUNX1* and *BCOR* identified patients unlikely to achieve sustained hematologic improvement, suggesting a secondary block to differentiation. These findings support the integration of mutational and molecular profiling to refine patient selection and guide combinatorial strategies in *IDH*-mutated myeloid malignancies.

**Key Points:** • Distinct molecular response patterns accompany hematologic benefit following IDH1 versus IDH2 inhibition.

• RUNX1 and BCOR mutations predict inferior hematologic response and shorter duration of IDH inhibitor therapy.

## Introduction

Mutations in *IDH1* and *IDH2* are recurrent in myeloid malignancies and result in neomorphic enzyme activity that produces 2-hydroxyglutarate, leading to epigenetic dysregulation and impaired hematopoietic differentiation ^1, 2, 3^. Recurrent oncogenic *IDH* mutations in myeloid neoplasms occur at the R132 locus of IDH1 and R140 or R172 residues of IDH2 ^1^.

Pharmacologic inhibition of mutant IDH enzymes with ivosidenib (*IDH1*) or enasidenib (*IDH2*) reduces 2-hydroxyglutarate levels and promotes differentiation, establishing these agents as a paradigm of differentiation therapy in acute myeloid leukemia (AML) ^4, 5^. Unlike cytotoxic therapies, the clinical activity of IDH inhibitors is characterized not only by remission induction but also by hematologic improvement, including recovery of neutrophil and platelet counts and reduction in transfusion burden, which may have meaningful benefits through enhanced quality of life and improved clinical outcomes ^6, 7^. These hematologic benefits may occur independently of complete morphologic remission, suggesting that therapeutic response to IDH inhibition reflects relief of differentiation block rather than eradication of mutant clones. However, only a subset of patients derive durable hematologic benefit and the biological determinants of response remain incompletely understood.

Emerging evidence suggests that persistence of mutant IDH clones during therapy is common, raising the possibility that clinical response may be partially decoupled from clonal clearance ^8^. Furthermore, co-occurring mutations in transcriptional and epigenetic regulators may impose a secondary block to differentiation, thereby limiting the efficacy of IDH inhibition. In particular, such classes of mutations may constrain lineage maturation even in the presence of effective IDH inhibition.

In addition, whether there are biologically meaningful differences between IDH1 and IDH2 inhibition remains unclear. Although both agents target related metabolic pathways, differences in clinical outcomes and molecular responses across studies raise the possibility of distinct mechanisms of action or differential capacity to induce clonal contraction.

In this study, we performed a retrospective analysis of patients with IDH-mutant myeloid malignancies treated with ivosidenib or enasidenib to define molecular predictors of hematologic response. We specifically investigated (1) changes in *IDH* variant allele frequency (VAF) as a measure of clonal dynamics, (2) differences between IDH1 and IDH2 inhibition, and (3) the impact of co-occurring mutations on hematologic response and treatment durability. Through this approach, we sought to identify biological features associated with response versus resistance to IDH inhibitor therapy and to inform strategies for patient selection and combination treatment.

## Methods

### Patient Selection

The study was approved by institutional review butaoards at Columbia University Irving Medical Center (IRB AAAU4817). We searched all adult patients with a hematological malignancy treated from 2010 through May 2025. Patients were included in the study if they had documented evidence of either an *IDH1* or *IDH2* mutation and were confirmed to have received either ivosidenib or enasidenib (Figure 1A). The presence of an *IDH* mutation was confirmed through chart review of next-generation sequencing mutational profiles using a targeted panel with a minimum sequencing depth of 500x and variant allele frequency (VAF) detection threshold of 2%.

**Figure 1.**
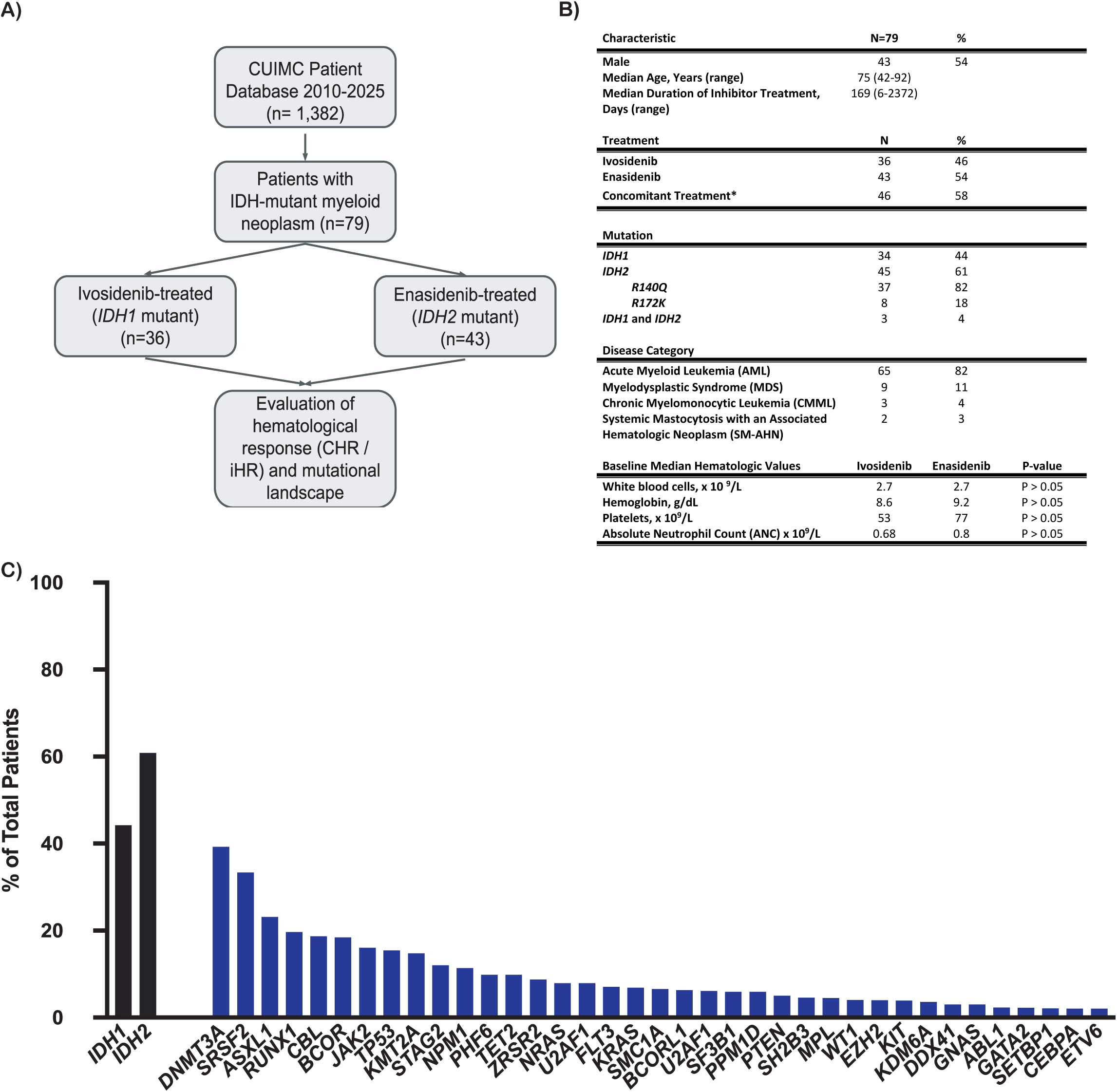
Patient Cohort and Baseline Genomic Landscape. (A) Cohort of patients selected from data repository. Eligible patients were those confirmed to have *IDH* mutations on molecular profiling and had received IDH1/2 inhibitor therapy. (B) Baseline patient demographics, clinical characteristics, and hematologic parameters. Asterisk denotes treatment with concomitant therapy which included: hydroxyurea, decitabine, azacytidine, and venetoclax. (C) Mutational landscape of total patient cohort at initiation of therapy. Bars are representative of frequencies of co-mutated genes, calculated as the number of patients who tested positive for the mutation, divided by the total number of patients tested for each specific mutation.

### Data Collection

Data collected included baseline hematologic laboratory values, time frame of red blood cell transfusions, demographics, mutational profiles, duration of treatment, and reasons for therapy discontinuation. The data collection end date was May 31st, 2025. Data was retrieved through retrospective chart review at three different timepoints if available - immediately prior to initiation of therapy, during the period of best response, and at time of discontinuation. Treatment duration was determined as the start of therapy until the date of therapy discontinuation or last follow-up. We documented whether patients received IDH inhibitors as monotherapy or in combination with other agents, including hypomethylating agents and venetoclax.

The cytogenetic risk category of AML and MDS patients was determined based on the 2022 European LeukemiaNet (ELN) risk classification and IPSS-R cytogenetic risk groups respectively ^9,10^. Gene mutation frequency was calculated as the number of patients who tested positive for the mutation, divided by the total number of patients tested for that particular mutation. Time to discontinuation of therapy was defined as the interval from IDH inhibitor therapy initiation to treatment discontinuation for any reason, including death, progression of disease, or treatment-related adverse events.

### Determination of Hematologic Response and Changes in Variant Allele Frequency

Hematologic response parameters were defined as absolute neutrophil count (ANC) ≥1.0 x 10^9^/L, platelet count ≥100 x 10^9^/L (as per 2022 ELN AML response criteria ^9^, and red blood cell transfusion independence, defined as the absence of any transfusions for ≥12 weeks within the first 24 weeks of receiving therapy ^11^. Patients achieving all 3 hematologic parameters were considered to have a complete hematologic response (CHR) and those with 0, 1, or 2 parameters were defined as having an incomplete hematologic response (iHR). CHR is distinct from ELN-defined CR/CRi and was designed to capture hematologic benefits independent of blast clearance. For AML patients, bone marrow blast percentage reduction to <5% was also assessed but not included in the primary CHR definition.

Variant allele frequency (VAF) of *IDH* mutations was compared at initiation of inhibitor therapy and at time of best response. Time of best response was determined as the time period during which all hematologic criteria were met. For patients with dual *IDH1*/*2* mutations (4% of cohort), we analyzed changes in both mutations separately.

### Sequencing

Clinical sequencing data for patients treated with IDH inhibitors were obtained from the electronic medical record in a de-identified manner under Institutional Review Board approval (AAAU4817). Genomic profiling was performed using a custom targeted next-generation sequencing panel (Archer; Integrated DNA Technologies) through the Columbia University Irving Medical Center Laboratory of Personalized Genomic Medicine. The test was performed in a College of American Pathologists–accredited and Clinical Laboratory Improvement Amendments–approved laboratory and was approved by the Clinical Laboratory Evaluation Program of the New York State Department of Health. This assay interrogates recurrently mutated genes in myeloid malignancies with a validated limit of detection of 2.7% variant allele frequency (VAF) ^12^.

For additional timepoints not captured through routine clinical testing, bone marrow aspirate and/or peripheral blood samples were retrieved from patients treated at Columbia University Irving Medical Center following written informed consent under a separate Institutional Review Board–approved protocol (AAAS7370), in accordance with the Declaration of Helsinki. Sequencing was performed at baseline prior to initiation of IDH inhibitor therapy and, when available, at serial timepoints, including best hematologic response and/or treatment discontinuation.

Variant calling was conducted using standardized clinical pipelines, and only pathogenic or likely pathogenic variants were included in downstream analyses. Variant allele frequencies were used to quantify clonal burden and assess longitudinal changes over the course of therapy, including dynamics of *IDH* mutations and the expansion or contraction of cooccurring clones.

Patients from each IDH inhibitor cohort with evaluable clinical outcomes (CHR or iHR) were selected for serial analysis when longitudinal samples were available. Patients with sequencing data available at multiple timepoints were selected for fish plot creation, to illustrate patterns of clonal evolution and changes in mutation burden throughout disease course following IDH inhibition.

### Statistical Analysis

Variant allele frequency (VAF) comparisons at treatment initiation and hematologic response were assessed using paired t-tests and changes in mean VAF between IDH inhibitor groups were assessed using the Mann-Whitney U test. Differences in co-mutation burden were analyzed by Fisher’s exact test with 2 x 2 tables, from which odds ratios and 95% confidence intervals were derived. Kaplan-Meier survival curves were used to estimate probabilities of therapy continuation and illustrate time to treatment discontinuation, for each inhibitor and co-mutational cohort. Differences in Kaplan-Meier curves were assessed using log-rank tests.

Multivariate analysis using Cox proportional hazards regression was performed to adjust for potential confounding factors including age, disease status, prior therapies, and concomitant treatments when assessing the impact of co-mutations on response and treatment duration. For multiple hypothesis testing when analyzing co-mutation patterns, we applied the Benjamini-Hochberg procedure to control the false discovery rate. All statistical tests were two-sided with a significance threshold of α = 0.05. All statistical analyses were performed using GraphPad Prism version 10 and R version 4.4.1.

## Results

### Patient Characteristics

Baseline patient characteristics are illustrated in Figure 1B. A total of 79 patients were identified as having *IDH* mutations and were treated with an IDH inhibitor. Median age was 75 years (range 42-92) and 54% of the cohort was male. At diagnosis, sixty-five patients had AML (82%), 9 (11%) had MDS, 3 (4%) had CMML, and 2 patients (3%) were diagnosed with systemic mastocytosis with an associated hematologic neoplasm (SM-AHN). The median follow-up time for the total cohort was 11.1 months (range 0.4 - 120 months).

Of patients with evaluable AML cytogenetics, based on ELN 2022 criteria, 5 (8%) patients had favorable risk, 23 patients (37%) had intermediate, and 34 (55%) had adverse risk. In accordance with the IPSS-R cytogenetic risk group, all MDS patients were characterized as having good cytogenetics.

Thirty four patients (44%) were *IDH1* mutated, 45 (61%) were *IDH2* mutated, and 3 (4%) had concurrent *IDH1* and *IDH2* mutations. Of *IDH2*-mutant patients, 37 (82%) possessed mutations at R140Q and 8 (18%) at R172K. All *IDH1*-mutant patients had mutations at codon R132. Between enasidenib and ivosidenib cohorts, there were no significant differences in baseline starting white blood cell count (WBC), ANC, hemoglobin, or platelet count (Figure 1B).

### IDH Inhibitor Therapy and Pharmacologic Data

Thirty-six patients (46%) were treated with ivosidenib and 43 (54%) with enasidenib (Figure 1B). Median duration of IDH inhibitor therapy was 169 days (range 6–2372). Forty-six patients (58%) received concomitant therapy, which included hydroxyurea, decitabine, azacytidine, and venetoclax. Twenty-four (30%) patients were treatment-naive and were treated in the frontline setting and 38 patients (48%) had previously received a hypomethylating agent such as azacitidine or decitabine.

The starting dosage of ivosidenib was 500 mg, with the exception of one patient receiving 250 mg. Ivosidenib was dose-reduced to 250 mg in 6 patients, primarily due to drug interactions with anti-fungal/azole agents. In the majority of patients, the starting dose of enasidenib was 100 mg, except for one patient who started at 200 mg (who was enrolled in AG-221 trial). In two patients, enasidenib was dose reduced to 50 mg due to adverse effects (hyperbilirubinemia).

### Molecular Landscape of Cohort at Initiation of Therapy

Among the total cohort of patients at initiation of IDH therapy, the most common co-occurring mutations at treatment initiation were *DNMT3A* (39%), *SRSF2* (33%), *ASXL1* (23%), *RUNX1* (20%), *CBL* (18%), and *BCOR* (18%) (Figure 1C). There was an average of 3 mutated genes per patient. CHR and iHR patients harbored a median mutation burden of 4 and 5 mutations respectively.

Of ivosidenib-treated patients with evaluable response data (n= 18), 5 patients (28%) cleared the *IDH1* mutation during the hematologic response period, with 4 of these patients achieving CHR. The remaining 13 patients had persistent IDH1 mutations. During enasidenib therapy, only 2 patients (9%) cleared the *IDH2* mutation and 19 possessed persistent *IDH2* mutations. All three patients with concurrent *IDH1*/*IDH2* mutations were treated with ivosidenib and only achieved iHR.

### Assessment of Hematologic Response

Overall, 34 patients (43%) achieved complete hematological response (CHR) (Figure 2A). This response rate aligns with published data showing a clinical objective response rate of 39-46% in relapsed/refractory settings ^13^. Of patients achieving CHR, 21 (62%) were treated with enasidenib and 13 (38%) with ivosidenib. There was no significant difference in the rates of CHR between IDH inhibitors (p>0.05). There was no significant association of rates of CHR with baseline demographic characteristics including age, gender, and prior hypomethylating agent exposure (Supplemental Figure 1).

**Figure 2.**
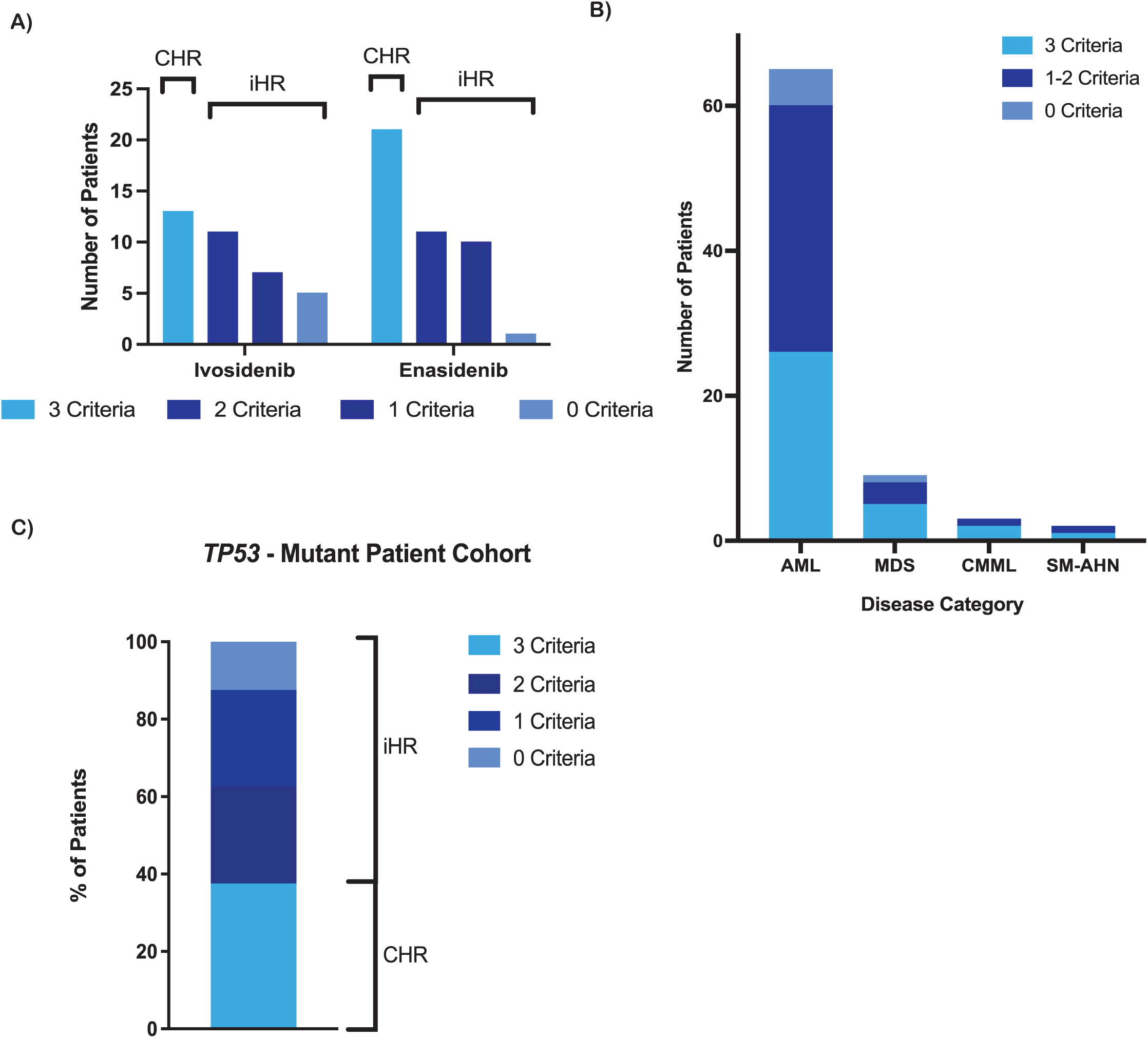
Hematological Response to IDH Inhibition. (A) Number of patients achieving complete hematologic response (CHR) or incomplete hematologic response (iHR) following therapy with an IDH inhibitor. (B) Hematologic response criteria achieved by each disease category. AML: acute myeloid leukemia, MDS: myelodysplastic syndrome, CMML: chronic myelomonocytic leukemia, SM-AHN: systemic mastocytosis with an associated hematologic neoplasm. (C) Distribution of hematologic criteria met across the *TP53*-mutated patient cohort (n = 8).

When stratified by disease type, CHR was achieved in 28/65 (43%) of AML patients, 4/9 (44%) of MDS patients, 1/3 (33%) of CMML patients, and 1/2 (50%) of SM-AHN patients, with no statistically significant differences between disease groups (p=0.96) (Figure 2B). Forty-five patients (57%) had incomplete hematologic response (iHR), with 22 patients meeting 2 criteria, 17 meeting one criteria, and 6 patients meeting no criteria. Of patients with iHR, 22 (49%) were treated with enasidenib and 23 (51%) with ivosidenib.

There were a total of 8 patients harboring *TP53* mutations and 3 (37.5%) achieved CHR while 5 patients (62.5%) had iHR (Figure 2C). With regards to individual hematologic parameters, among the *TP53 -* mutated patients, 5 (63%) demonstrated platelet response, 6 (75%) showed ANC recovery, and 3 (38%) had transfusion independence.

### Changes in Variant Allele Frequency During Hematologic Response

Baseline mean VAF at treatment initiation was not significantly different between CHR vs. iHR among either IDH-inhibitor group (*p* >0.05). Patients receiving ivosidenib who achieved CHR demonstrated a significant reduction in *IDH1* VAF during hematologic response (*p* = 0.010) (Figure 3A). In contrast, enasidenib-treated patients achieving CHR did not show a significant change in *IDH2* VAF (*p* > 0.05) (Figure 3A). No difference in *IDH* VAF was observed in patients achieving iHR in either inhibitor group (*p* > 0.05) (Figure 3A). When comparing the magnitude of *IDH* VAF changes among CHR patients, ivosidenib-treated patients showed a significantly greater reduction in VAF as compared to patients who received enasidenib (mean VAF change -16.73 vs -0.05, *p = 0.034*) (Figure 3B).

**Figure 3.**
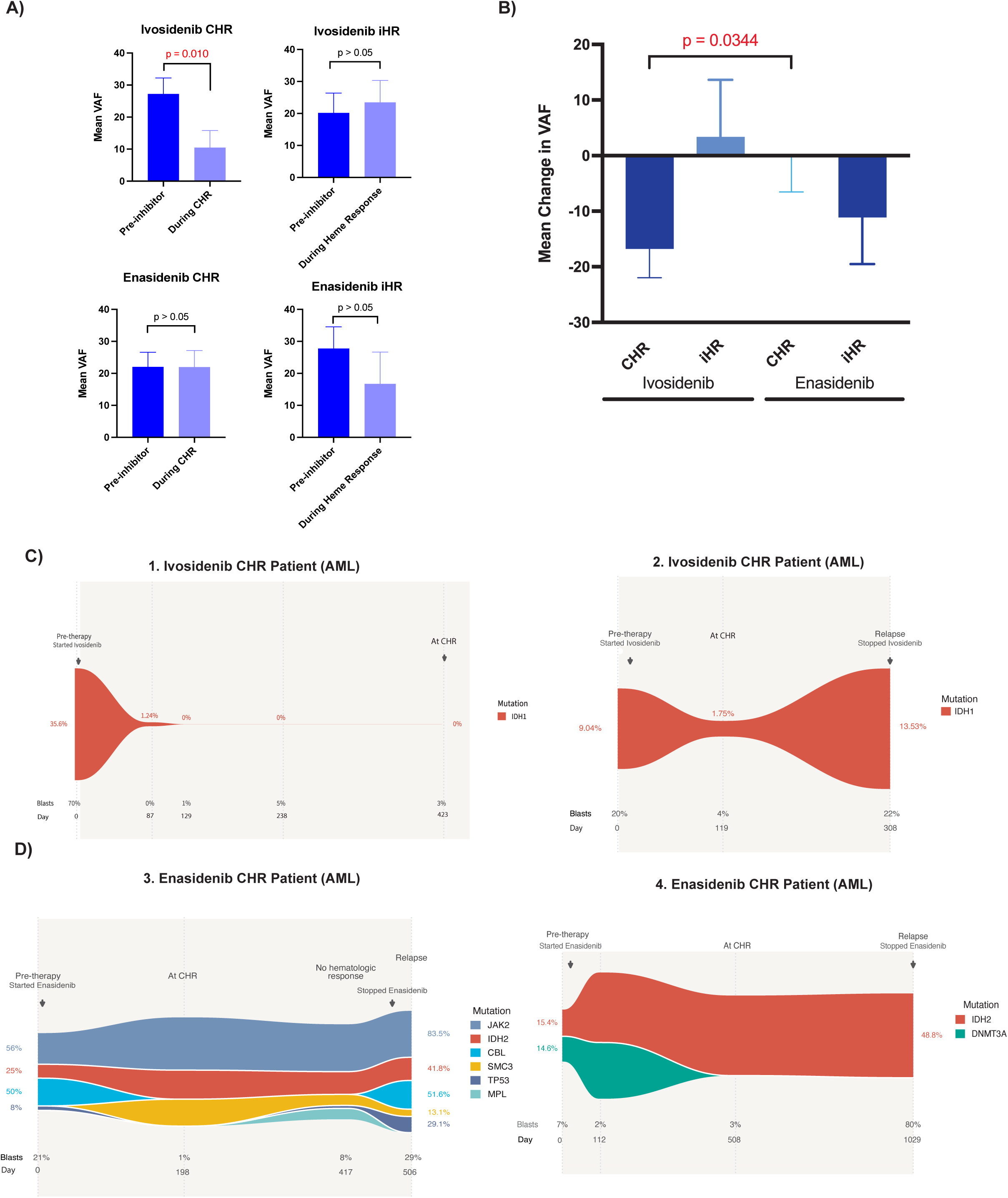
Differences in Molecular Response Between IDH Inhibitors and Longitudinal VAF Trajectories. (A) Bar graphs comparing mean *IDH* variant allele frequency (VAF), for each inhibitor and response group, at two timepoints: prior to inhibitor and during best hematologic response. Error bars represent standard error of the mean and *p -* values are labeled for each comparison group. (B) Bar graph illustrating comparison of changes in mean *IDH* VAF, among patients receiving each inhibitor and response category. Ivosidenib CHR patients had significantly greater reduction in VAF relative to the enasidenib CHR cohort (*p* = 0.034). (C) Longitudinal mutational dynamics of *IDH1* in an ivosidenib-treated patient with AML. In both patients, only *IDH1* molecular testing was performed. *IDH1* mutation VAF is shown in parallel with corresponding bone marrow blast percentages with corresponding times of inhibitor initiation and discontinuation. X-axis is scaled to reflect time (in days) elapsed between timepoints. (D) Serial molecular profiling of two representative *IDH2-*mutant AML patients during enasidenib therapy. Molecular testing was performed using a comprehensive targeting mutational panel. Bone marrow blast percentages and disease status are shown for each timepoint.

Serial molecular profiling for an *IDH1-*mutated AML patient treated with ivosidenib showed rapid molecular clearance of the *IDH1* mutation after initiation of therapy, which persisted until the hematologic response period (Figure 3C). In another patient with *IDH1*-mutant AML, following ivosidenib exposure, there was clonal contraction of the *IDH1* mutation observed during CHR, with subsequent molecular expansion during disease relapse (Figure 3C). Serial molecular testing for two enasidenib-treated AML patients achieving CHR, demonstrated stable *IDH2* VAF despite blast clearance and hematologic improvement, with persistent mutational burden through hematologic response and disease relapse (Figure 3D). The baseline demographic, clinical, and cytogenetic characteristics of the illustrated patients are shown in Supplemental Table 1.

### Association of Co-mutated Genes with Hematologic Response and Therapy Duration

A significantly greater proportion of co-mutated *RUNX1* patients were associated with iHR (*p*=0.0016) and *TET2* co-mutations trended towards significance (*p* = 0.069) (Figure 4A, Supplemental Figure 2). Notably, none of the patients achieving CHR were found to have *RUNX1* or *TET2* mutations (0/23 and 0/21 respectively) (Figure 4A). This finding aligns with previous reports identifying *RUNX1* and *TET2* mutations as potential mechanisms of resistance to IDH inhibitors (^14^).

**Figure 4.**
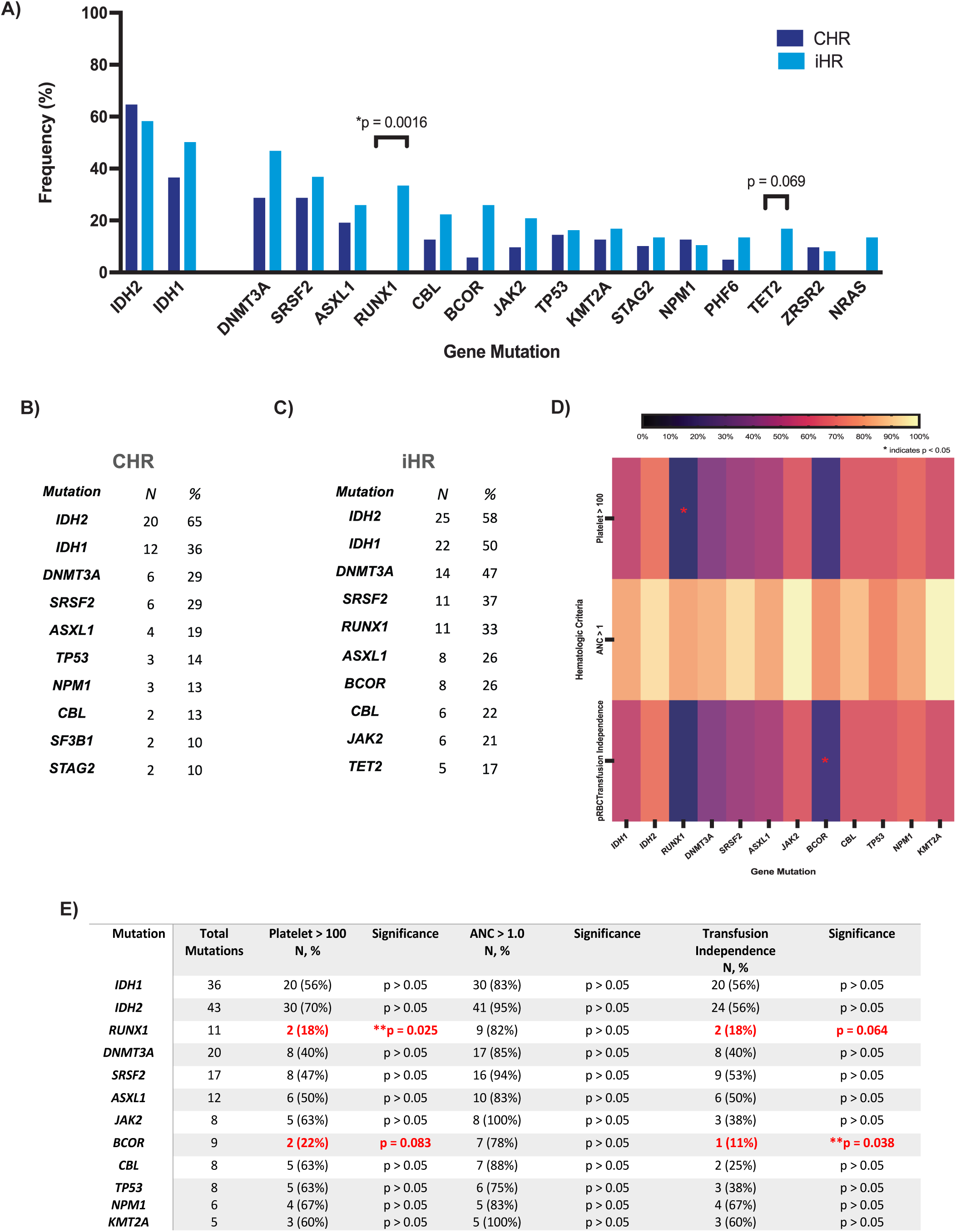
Co-mutational Landscape Predicts Failure of Hematological Response. (A) Frequencies of co-mutated genes at initiation of therapy, in patients achieving CHR vs iHR. *RUNX1* mutations were significantly enriched in the iHR cohort (*p =* 0.0016) and *TET2* mutations trended towards significance (*p =* 0.069). (B) Most commonly mutated genes in the CHR cohort were *DNMT3A, SRSF2,* and *ASXL1*. (C) The most frequently co-mutated genes in the iHR group were *DNMT3A, SRSF2,* and *RUNX1*. (D and E) Association of recurrent mutations with individual hematologic response criteria (red blood cell transfusion independence, absolute neutrophil count (ANC), platelet count). Red asterisks denote statistically significant associations between specific gene mutations and response criteria (p < 0.05). *RUNX1* and *BCOR* mutations were associated with inferior platelet response and red blood cell transfusion independence respectively (*p =* 0.025, *p =* 0.038).

Within patients achieving CHR, the most commonly co-mutated genes were *DNMT3A* (29%), *SRSF2* (29%), and *ASXL1* (19%) (Figure 4B). In the iHR cohort, most common co-mutations included *DNMT3A* (47%), *SRSF2* (37%), *RUNX1* (33%), *ASXL1* (26%), and *BCOR* (26%) (Figure 4C).

When stratified by hematologic criteria, *RUNX1* and *BCOR* mutations were observed to have the lowest rates of meeting platelet count and transfusion independence parameters. A significantly lower proportion of *RUNX1* mutated patients sustained platelet recovery (*p* = 0.025, 95% CI 0.018 - 0.88, OR 0.18) (Figure 4D, 4E). Furthermore, a lower proportion of *RUNX1* mutated patients achieved transfusion independence, although this finding approached significance (*p* = 0.064, 95% CI 0.024 - 1.19, OR 0.24). Similarly, *BCOR* co-mutated patients demonstrated significantly lower rates of red blood cell transfusion independence (*p* = 0.038, 95% CI 0.003 - 1.032, OR 0.14) (Figure 4D, 4E).

Using Kaplan Meier curves to assess time to treatment discontinuation for each co-mutation, *RUNX1*-mutated patients showed a significantly greater probability of terminating therapy relative to non-mutated patients (hazard ratio [HR] 2.84, 95% CI 1.09-7.40, *p* = 0.0017) (Figure 5C). In *RUNX1-*mutated patients, therapy was most frequently discontinued due to transition to hospice or comfort focused measures (45%). Furthermore, *BCOR-*mutated patients were significantly more likely to discontinue therapy relative to non-mutated individuals (HR 2.68, 95% CI 0.89-8.09, *p* = 0.0097) (Figure 5D). Among the *BCOR-*mutated patients, 56% discontinued therapy due to transitions to hospice, progression of disease, or adverse effects. Comprehensive analysis of therapy duration for additional genetic co-mutations is shown in Supplemental Figure 4.

**Figure 5.**
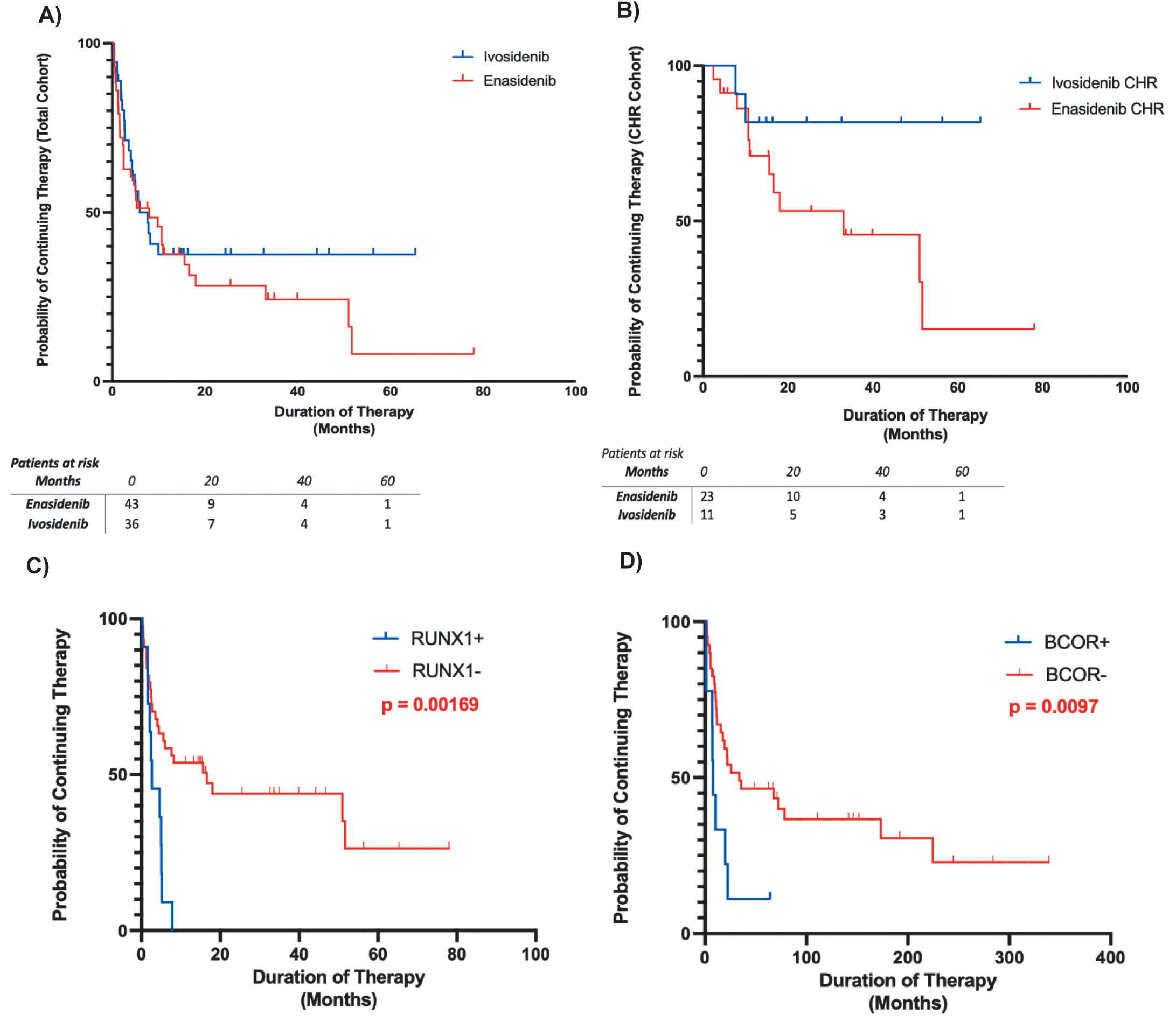
Durable Treatment Benefit is Mutation-Dependent. (A) Kaplan-Meier curve illustrating time to treatment discontinuation among all patients receiving ivosidenib or enasidenib. Time to treatment discontinuation was defined as interval from IDH inhibitor therapy initiation to treatment discontinuation for any reason. There was no significant difference in duration of therapy between the IDH inhibitor groups (*p =* 0.409). (B) Kaplan-Meier graph depicting time to treatment discontinuation among all patients with CHR, stratified by IDH inhibitor. Enasidenib-treated CHR patients had greater probability of discontinuing therapy relative to the ivosidenib CHR cohort (*p =* 0.0908). (C) Kaplan-Meier analysis of time to treatment discontinuation, as stratified by *RUNX1* mutational status. Patients with *RUNX1* co-mutations (n = 11) had significantly greater probability of discontinuing therapy as compared to wildtype patients (HR 2.84, *p =* 0.0017). (D) Kaplan-Meier curve illustrating time to treatment discontinuation, stratified by *BCOR* co-mutational status. *BCOR-*mutational status (n = 8) was associated with shorter treatment duration compared to non-mutated patients (HR 2.31, *p□*=□0.0058).

In multivariate analysis adjusting for age, disease status, prior therapies, and concomitant treatments, both *RUNX1* (adjusted HR 2.53, 95% CI 1.42-4.51, *p* = 0.0024) and *BCOR* mutations (adjusted HR 2.31, 95% CI 1.28-4.17, *p*=0.0058) remained independently associated with shorter treatment duration, suggesting their role as potential biomarkers of primary resistance to IDH inhibition.

### IDH Inhibitor Treatment Duration and Time to Treatment Discontinuation

Overall, IDH therapy was discontinued in 54 patients (68%) with five patients terminating therapy to undergo hematopoietic stem cell transplantation. Other reasons for discontinuation of therapy included progression of disease, death, transition to hospice or comfort-focused measures, and adverse effects including hyperbilirubinemia or anorexia. Therapy was discontinued in one patient due to differentiation syndrome. This incidence is lower than the 19-31% reported in clinical trials and real-world studies ^15^ and likely represents prophylactic measures or differences in monitoring.

There was no significant difference in treatment duration between the two IDH inhibitor groups (*p* = 0.409) (Figure 5A). There was also no difference in time to therapy discontinuation between patients with and without prior hypomethylating agent exposure (*p =* 0.36, Supplemental Figure 3). When stratified by patients with CHR, enasidenib patients had greater probability of discontinuing therapy than the ivosidenib cohort, and this difference approached significance (*p* = 0.0908) (Figure 5B). This finding suggests that ivosidenib might provide more durable responses in patients who achieve initial hematologic improvement, consistent with the greater VAF reduction observed in ivosidenib responders.

## Discussion

IDH inhibitors represent a novel therapeutic class that targets the epigenetic dysregulation underlying impaired differentiation in myeloid malignancies. By reversing the accumulation of 2-hydroxyglutarate and restoring TET and histone demethylase activity, these agents relieve the differentiation block characteristic of IDH-mutant myeloid malignancies ^16^. In this study, we provide real-world evidence that response to IDH inhibition is characterized by heterogeneous patterns of clonal dynamics and is strongly influenced by co-mutational context. As differentiation therapies may improve hematopoiesis without inducing morphologic remission, CHR was selected to capture clinically meaningful hematologic benefit independent of blast clearance. Our findings support a model in which hematologic improvement reflects relief of differentiation block rather than complete eradication of the leukemic clone, and in which secondary genetic lesions may limit the durability and depth of response.

A key observation from our analysis is the differential molecular response observed between IDH1 and IDH2 inhibition. Among patients treated with ivosidenib, achievement of complete hematologic response (CHR) was associated with a greater reduction in *IDH1* VAF, consistent with measurable clonal contraction.

In contrast, enasidenib-treated patients did not demonstrate a comparable association between *IDH2* VAF reduction and hematologic response. This pattern is consistent with a differential capacity of IDH1 versus IDH2 inhibition to induce clonal suppression, which may contribute to differences in treatment durability. Prior studies have similarly suggested higher rates of molecular response and hematologic improvement with IDH1-directed therapy, supporting the biological distinction observed in our cohort ^5,8,17^.

Importantly, our findings highlight that hematologic response and clonal dynamics are not uniformly coupled. While IDH1 inhibition was associated with measurable reductions in mutant allele burden in responders, many patients—particularly those treated with enasidenib— demonstrated hematologic improvement without substantial VAF decline. These observations reinforce the notion that differentiation-based therapies may decouple clinical benefit from molecular clearance, distinguishing them from cytotoxic approaches ^18^. At the same time, the greater degree of clonal contraction observed in ivosidenib-treated responders raises the possibility that combined differentiation and clonal suppression may contribute to more durable benefit.

Co-mutational context emerged as a key determinant of response. Mutations in *RUNX1* and *BCOR* were strongly associated with incomplete hematologic response and shorter duration of therapy. No patients harboring *RUNX1* mutations achieved complete hematologic recovery. These findings support a model in which alterations in transcriptional and epigenetic regulators impose a secondary block to differentiation downstream of IDH inhibition ^19^. Notably, none of the patients achieving CHR harbored *RUNX1* or *TET2* mutations, further suggesting a constraint on differentiation in these molecular subsets. Given the role of RUNX1 in hematopoietic lineage specification and BCOR in chromatin remodeling through the PRC1.1 complex, disruption of these pathways may limit the ability of IDH inhibition to fully restore normal maturation programs ^20^. This concept is supported by prior studies demonstrating the emergence of mutations in transcriptional regulators at relapse following IDH inhibitor therapy, implicating these alterations in both primary and acquired resistance ^14^.

From a clinical perspective, these results have several implications. First, baseline mutational profiling may help identify patients less likely to derive meaningful hematologic benefit from IDH inhibitor monotherapy, particularly those harboring *RUNX1* or *BCOR* mutations. For patients harboring these high-risk mutations, rational combination strategies warrant investigation. The triplet regimen of ivosidenib with venetoclax and azacitidine has shown promising efficacy with 90% composite complete remission rates and appears to overcome established resistance mechanisms to single-agent IDH inhibition ^21^. Similarly, newer irreversible IDH inhibitors such as LY3410738 may overcome resistance mutations that impair binding of first-generation inhibitors ^22^.

Second, longitudinal molecular monitoring of IDH VAF may provide complementary information regarding treatment depth and durability, particularly in patients treated with IDH1-directed therapy. Together, these findings support a more integrated approach to therapeutic decision-making that incorporates both genomic context and molecular response kinetics. Both next-generation sequencing and droplet digital PCR have demonstrated utility for IDH1/2 minimal residual disease monitoring ^23^, potentially allowing for early detection of emerging resistant clones and timely therapeutic intervention. Longitudinal assessment of VAF dynamics, particularly in patients with *RUNX1* or *BCOR* co-mutations, may reveal patterns of clonal selection under therapeutic pressure and inform optimal treatment duration or sequencing. Ultimately, integration of genomic profiling with clinical parameters could lead to personalized approaches that maximize response durability while minimizing toxicity in this molecularly heterogeneous patient population.

We acknowledge that this study has several limitations. The retrospective design and modest sample size limit the ability to draw definitive conclusions regarding causality, and the inclusion of heterogeneous disease subtypes (AML, MDS, CMML, and SM-AHN) may introduce confounding, yet similar response rates across groups. Molecular testing was not uniformly performed at all timepoints, and the number of patients with paired longitudinal sequencing was limited. Additionally, the relatively high proportion of patients receiving concomitant therapy represents a potential source of confounding, although multivariate analyses were performed to adjust for clinical factors. Finally, our definition of complete hematologic response, while designed to capture clinically meaningful improvement, is distinct from standard response criteria and does not incorporate blast clearance. Prospective validation in larger, uniformly profiled cohorts will be necessary to confirm these findings. Despite these limitations, our findings underscore the real-world clinical benefit of IDH inhibition. Reductions in transfusion burden and improvements in hematologic parameters may translate into enhanced quality of life, particularly in patients with MDS ^24,25^.

In summary, our findings support a model in which the clinical activity of IDH inhibitors reflects relief of differentiation block, variably accompanied by clonal contraction, and modulated by co-occurring genetic alterations. Differences between IDH1 and IDH2 inhibition in their association with molecular response suggest distinct biological effects that may influence treatment durability. Co-mutations in *RUNX1* and *BCOR* identify a subset of patients with impaired hematologic recovery, consistent with a secondary block to differentiation. These results provide a framework for integrating genomic profiling and molecular monitoring into the management of *IDH*-mutated myeloid malignancies and underscore the need for combination strategies to overcome resistance in biologically defined subgroups.

## Data Availability

Data produced in the present study are available upon reasonable request to the authors.

## Acknowledgements

A.S. is supported by an R38 CAPRI. M.K. is Supported by a Young Investigator Award from the Edward P. Evans Foundation. B.C is supported by a Research Training award for Fellows by the American Society of Hematology and Young Investigator Award from the American Society of Clinical Oncology. V.S.S. is supported by an American Society of Hematology HONORS award and American Society for Clinical Investigation Physician-Scientist Support Foundation fellowship. A.D.V is supported by the Damon Runyon Cancer Research Foundation (CI-120-22), the Gabrielle’s Angels Foundation, and an American Society of Hematology Scholar Award.

## Disclosures

B.C is a scientific advisory board member for Cogent Biosciences, Sobi, Refined Oncology, Geron Corporation, PharmaEssentia and consultancy for Blueprint Medicines. A.D.V. is a scientific advisory board member of Arima Genomics and Pixelgen Technologies.

## Notes

### Competing Interest Statement

A.S. is supported by an R38 NIH/NCI physician-scientist award. M.K. is Supported by a Young Investigator Award from the Edward P. Evans Foundation. B.C is supported by a Research Training award for Fellows by the American Society of Hematology and Young Investigator Award from the American Society of Clinical Oncology. V.S.S. is supported by an American Society of Hematology HONORS award and American Society for Clinical Investigation Physician-Scientist Support Foundation fellowship. A.D.V is supported by the Damon Runyon Cancer Research Foundation (CI-120-22), the Gabrielle's Angels Foundation, and an American Society of Hematology Scholar Award. Disclosures: B.C is a scientific advisory board member for Cogent Biosciences, Sobi, Refined Oncology, Geron Corporation, PharmaEssentia and consultancy for Blueprint Medicines. A.D.V. is a scientific advisory board member of Arima Genomics and Pixelgen Technologies.

### Author Declarations

The study was approved by institutional review boards at Columbia University Irving Medical Center (IRB AAAU4817).

