## Supplemental Figures and Figure Legends for "Genetic predictors of clonal contraction and hematologic response in *IDH* mutant myeloid malignancies": Supp Figures 9_3.pdf

**Supplemental Figure 1.**

Association of Baseline Patient Demographics and Characteristics with Hematologic Response

| | Odds Ratio (iHR vs CHR) | P-value |
|---|---|---|
| **Age** | | |
| Age (in years) | 1.01 (0.96–1.07) | 0.637 |
| **Gender** | | |
| Male vs Female | 1.82 (0.60–5.70) | 0.291 |
| **Prior Hypomethylating Agent Exposure** | | |
| Yes vs No | 0.87 (0.28–2.74) | 0.809 |
| **Cytogenetics** | | |
| Adverse vs Intermediate | 5.90 (1.79–21.56) | 0.005 |
| Favorable vs Intermediate | 1.31 (0.13–11.37) | 0.807 |
| Good vs Intermediate | 1.42 (0.22–8.46) | 0.699 |

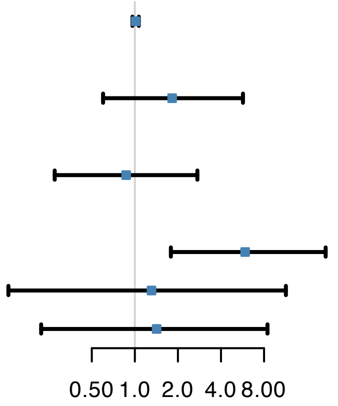


**Supplemental Figure 2.**

Association of Gene Co-Mutations with Hematologic Response

| Gene (CHR I iHR) | OR (iHR vs CHR) | P-value |
|---|---|---|
| *RUNX1** (CHR 0/23 I iHR 11/33) | Inf (2.23–Inf) | 0.002 |
| *TET2* (CHR 0/21 I iHR 5/30) | Inf (0.68–Inf) | 0.069 |
| *SH2B3* (CHR 2/16 I iHR 0/28) | 0.00 (0.00–2.98) | 0.127 |
| *BCOR* (CHR 1/18 I iHR 8/31) | 5.74 (0.66–277.13) | 0.127 |
| *NRAS* (CHR 0/21 I iHR 4/30) | Inf (0.48–Inf) | 0.134 |
| *DNMT3A* (CHR 6/21 I iHR 14/30) | 2.15 (0.58–8.75) | 0.250 |
| *ABL1* (CHR 1/15 I iHR 0/29) | 0.00 (0.00–20.17) | 0.341 |
| *PHF6* (CHR 1/21 I iHR 4/30) | 3.02 (0.27–158.84) | 0.391 |
| *ETV6* (CHR 1/20 I iHR 0/30) | 0.00 (0.00–26.00) | 0.400 |
| *GNAS* (CHR 1/14 I iHR 0/20) | 0.00 (0.00–27.30) | 0.412 |
| *JAK2* (CHR 2/21 I iHR 6/29) | 2.44 (0.38–27.41) | 0.441 |
| *EZH2* (CHR 0/21 I iHR 2/30) | Inf (0.13–Inf) | 0.506 |
| *KIT* (CHR 0/21 I iHR 2/31) | Inf (0.13–Inf) | 0.509 |
| *WT1* (CHR 0/20 I iHR 2/30) | Inf (0.12–Inf) | 0.510 |
| *SF3B1* (CHR 2/21 I iHR 1/30) | 0.33 (0.01–6.85) | 0.561 |
| *CBL* (CHR 2/16 I iHR 6/27) | 1.97 (0.29–22.69) | 0.688 |
| *ASXL1* (CHR 4/21 I iHR 8/31) | 1.47 (0.33–7.79) | 0.741 |
| *SRSF2* (CHR 6/21 I iHR 11/30) | 1.44 (0.38–5.91) | 0.763 |

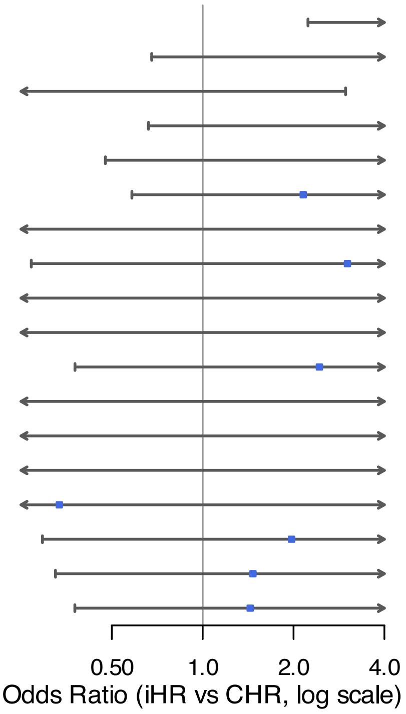


** Statistically significant (nominal p < 0.05)*

**Supplemental Figure 3.**

**Prior Hypomethylating Agent Exposure and Time to Treatment Discontinuation**
Hazard Ratio = 1.281    95% CI: 0.751 – 2.186

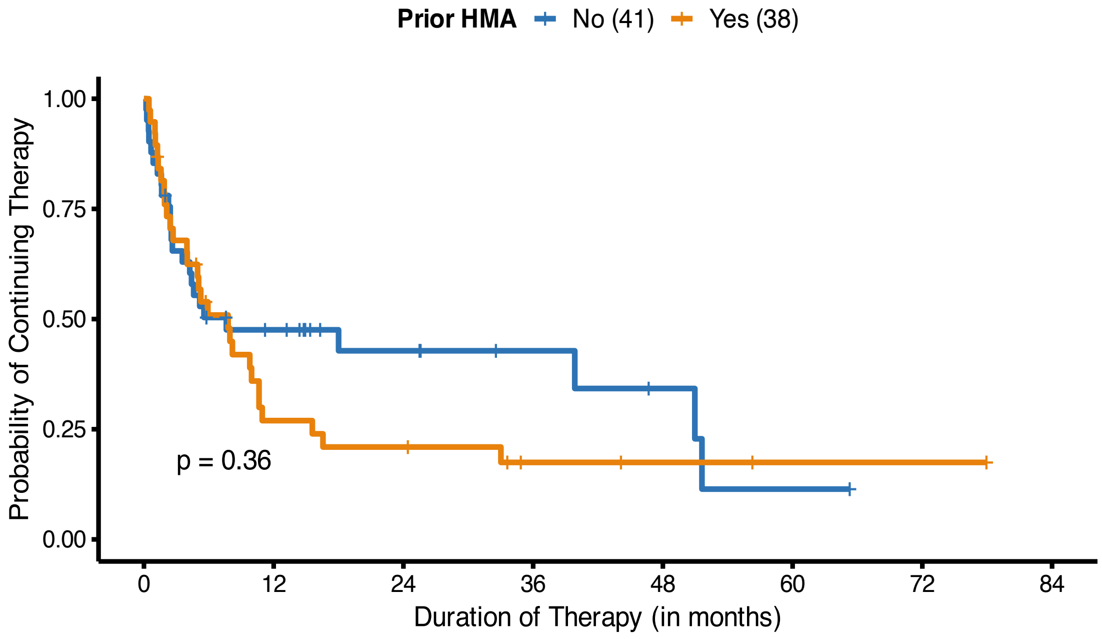


**Supplemental Figure 4.**

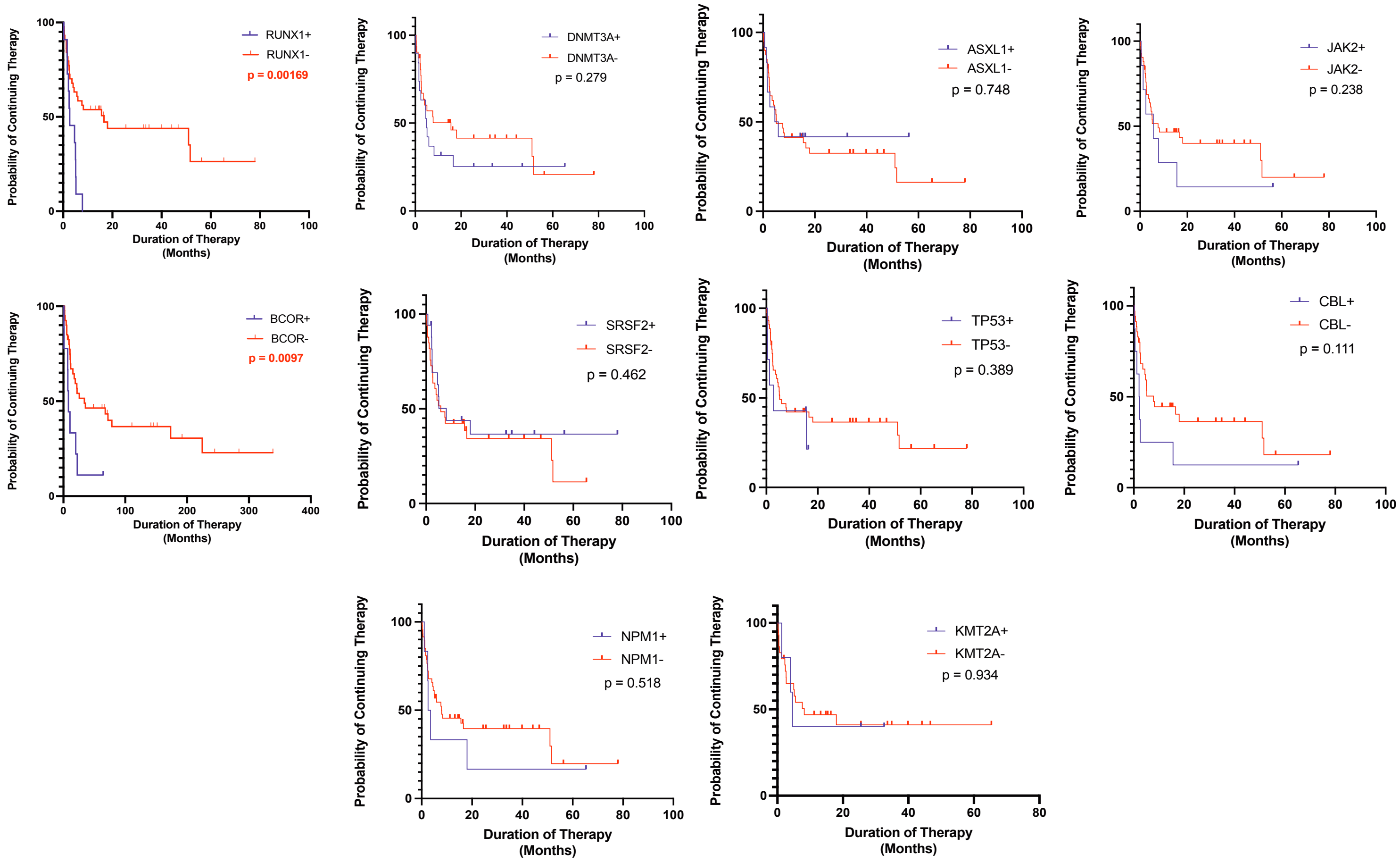

**Supplemental Figure 5.**

A)

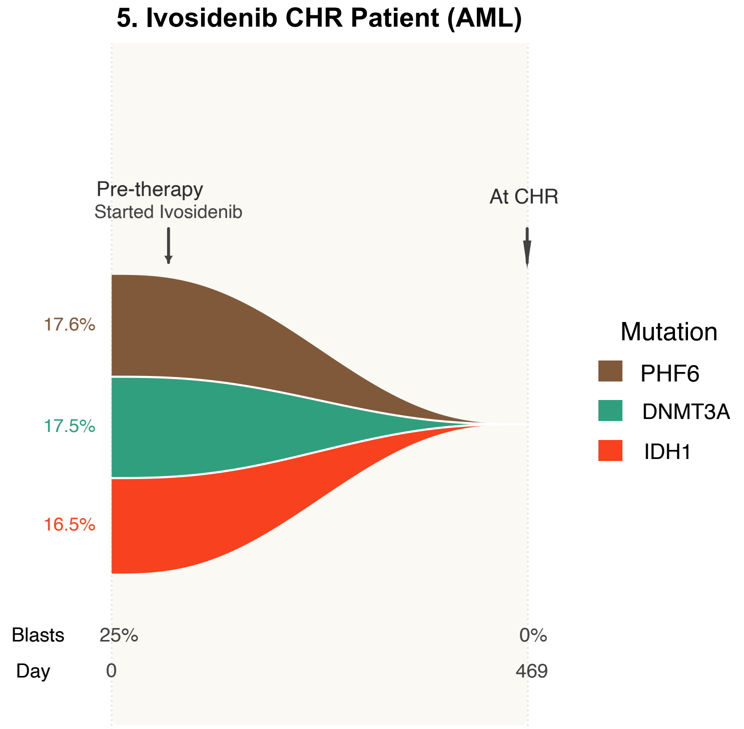


B)

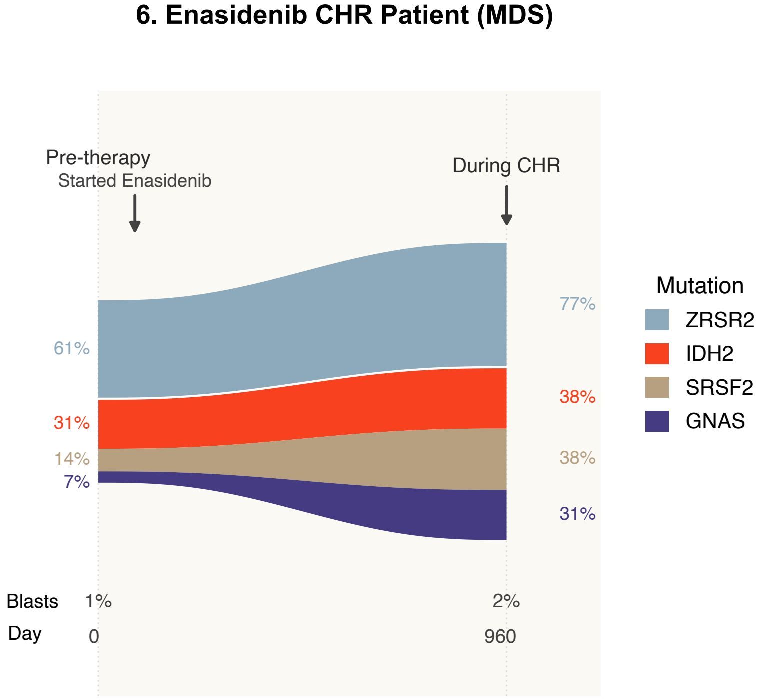


**Supplemental Table 1.**

| ID | Gender | Disease | Karyotype | Therapy | Mutations | Sequencing Panel | Specimen Type |
|---|---|---|---|---|---|---|---|
| **1** | Male | AML | 47,XY,+8[20] | Ivosidenib | *IDH1* | IDH only | Bone marrow |
| **2** | Male | AML | 47,XY,+8[1]/46,XY,add (10)(p13)[1]/46,XY[18] | Ivosidenib | *IDH1* | IDH only | Bone marrow |
| **3** | Female | AML | 46,XX,del(20)(q11.2) [1]/ 46,XX[18] | Enasidenib | *IDH2/TP53/JAK2/SMC3/CBL/ MPL* | Genotypix | Bone marrow |
| **4** | Female | AML | 47,XY[14]//46,XX[6] | Enasidenib | *IDH2/DNMT3A* | Neogenomics | Bone marrow |
| **5** | Female | AML | Normal | Ivosidenib | *IDH1/DNMT3A/PHF6* | Neogenomics | Bone marrow |
| **6** | Male | MDS | Normal | Enasidenib | *IDH2/SRSF2/GNAS/ZRSR2* | Targeted Myeloid Panel | Bone marrow |
