## Supplemental Figures and Figure Legends for "Genetic predictors of clonal contraction and hematologic response in *IDH* mutant myeloid malignancies": Supplemental_Figure_Legends_.docx

**Supplemental Figure 1**. Multivariate regression analysis to assess associations in achieving hematologic response with baseline patient demographics and characteristics, including age (in years), sex, prior hypomethylating agent (HMA) exposure, and cytogenetic classification.

**Supplemental Figure 2.** Forest plot of co-mutated genes and odds ratios correlating with association with hematologic response. *RUNX1* co-mutations were significantly associated with incomplete hematologic response (*p =* 0.002) while *TET2* mutations approached significance (*p =* 0.069).

**Supplemental Figure 3.** Kaplan-Meier analysis comparing time to IDH inhibitor treatment discontinuation among patients with and without prior hypomethylating agent exposure. There was no significant difference between the two groups (*p > 0.05*).

**Supplemental Figure 4.** Kaplan-Meier curves showing time to treatment discontinuation, as stratified by additional co-mutational genes.

**Supplemental Figure 5.** (A) Longitudinal molecular profiling of an *IDH1-*mutant AML patient achieving CHR with ivosidenib therapy. At CHR, the patient had clearance of the *IDH1, DNMT3A,* and *PHF6* mutations initially detected at initiation of IDH inhibitor therapy, which also correlated with blast clearance. Shown along the x-axis is correlating bone marrow blast percentage correlating to time (in days) from initial pre-treatment bone marrow sample. (B) Serial mutational profiling of an *IDH2*-mutant MDS patient who received enasidenib therapy. At CHR, *IDH2* VAF increased slightly from 31% to 38%.

**Supplemental Table 1**. Baseline demographic, clinical, and cytogenetic characteristics of patients selected to illustrate changes in clonal evolution and *IDH* variant allele frequency.
